# Evaluation of Protein Cards: A nutrition education tool for metabolic bariatric surgery

**DOI:** 10.1101/2025.01.31.25321467

**Authors:** Patricia F.C. Acosta, Alexandra J. Heidl, Patricia M. Angeles, Biagina-Carla Farnesi, Peggy Alcindor, Angela S. Alberga, Julius Erdstein, Stephanie Saputra, Tamara R. Cohen

## Abstract

**Background:** Metabolic bariatric surgery (MBS) is a safe, effective treatment for severe obesity and its associated comorbidities. However, adherence to postoperative guidelines, particularly dietary protein intake remains a challenge. This study examined the suitability of the *Protein Cards*, a protein-focused nutrition education tool developed to support individuals meet their protein requirements during the postoperative diet stages: fluid, purée, soft, and regular diets.

**Methods:** An online adapted version of the Suitability Assessment of Materials questionnaire was administered from September 2020 to May 2021. Participants were recruited via convenience sampling and advertisement. The tool was rated as “superior”, “adequate” or “not suitable” on content, literacy demand, graphic illustrations, layout and typography, learning stimulation and motivation, and cultural appropriateness, with scores of 2, 1, and 0, respectively.

**Results:** A total of 442 individuals completed the online survey. Participants were identified as individuals who have undergone MBS (n= 268), caregivers of individuals who completed MBS (n =68), and/or healthcare providers specializing in MBS (n =106). The *Protein Cards* received an overall “superior” rating of 73.16%. The tool had a high likelihood of use particularly for the soft diet stage (63.98 ± 20.71). Participants preferred the tool be available as a mobile application (63.46 ± 20.44) followed by paper book (59.42 ± 22.12) format.

**Conclusion:** The development of nutrition education tools is essential for supporting individuals who have undergone MBS in adopting healthy dietary habits, particularly in their meeting protein requirements. Future studies will refine the *Protein Cards* and evaluate its usability among individuals post-MBS.

## Introduction

Severe obesity characterized by a body mass index (BMI) ≥35 kg/m^2^ affects ∼1.9 million Canadian adults, [1] and is projected to double between 2020-2035. [2,3] This significantly impacts health, overall quality of life, [4] and healthcare costs. [5] While lifestyle interventions remain the first line of treatment for obesity, [6,7] metabolic bariatric surgery (MBS) has been found to be a safe and effective treatment option for severe obesity and its associated comorbidities [8]. Common procedures, such as vertical sleeve gastrectomy and Roux-en-Y gastric bypass, modify the gastrointestinal tract to significantly reduce food intake and capacity to facilitate reduction of excess body weight and lower associated health risks [9]. Notable postoperative benefits include sustained weight reduction, improved cardiometabolic health and health-related quality of life, and remission of type 2 diabetes mellitus [10–14].

The long-term success of MBS remains dependent on commitment to lifestyle changes, particularly, food choices and eating behaviors that significantly influence MBS outcomes. [15] However, non-adherence to dietary guidelines is frequently seen in the bariatric population [16–18]. Individuals report feeling overwhelmed [19] and uncertain about food choices, often consuming a single food item for weeks post-surgery. One way to increase adherence to postoperative dietary guidelines is by advancing through the various post-bariatric surgery diet stages: fluid, purée, soft, and regular diets [20,21]. The duration of each stage varies according to the patient’s tolerance, to ensure optimal healing of the gastrointestinal tract, minimize the risk of complications, and facilitate safe adaptation to foods [22].

One of the main nutritional concerns among patients post-MBS is inadequate dietary protein intake. [23,24] Protein intake has been demonstrated to be protective against the loss of fat-free mass and muscle mass, which can occur due to the rapid weight loss. [25] Postoperative protein intake has also been associated with increased satiety, weight loss, and improved body composition [25–27]. Current guidelines recommend 35% of total energy intake come from protein [28,29] or a minimum of 60 g/ day up to 1.5 g/kg ideal weight per day. [20,23,30,31] However, studies suggest that protein intake is often inadequate in this population, with a daily protein consumption of approximately 45 g [32,33]. In fact, protein deficiency remains the most severe macronutrient complication post-MBS, largely attributed to low intakes of protein-rich foods [23].

The use of nutrition education tools provide comprehensive and accessible information to increase knowledge and facilitate the adoption of healthy nutrition-related behaviors [34]. For these tools to be effective, they need be acceptable, suitable, and targeted to their end-users. However, there is a notable lack of tailored tools to help guide those who have undergone MBS in meeting their postoperative nutritional needs, particularly dietary protein intake. Majority of the materials have reading levels often exceeding what is appropriate for this demographic [35–39]. This is particularly concerning as health literacy was associated with successful weight loss among patients who have undergone MBS. [40,41]

As such, the *“Protein Cards”* tool was developed to facilitate meeting the nutritional needs post-MBS, with emphasis on protein-rich dietary choices. The objective of this study was to evaluate the suitability of a patient nutrition education tool called *Protein Cards* among individuals who have undergone MBS, their caregivers, and healthcare providers specializing in MBS.

## Materials and Methods

### Study Design

This study was a cross-sectional, online survey conducted from September 2020 to May 2021 to assess perceptions and feedback on the suitability of the *Protein Cards*. The self-administered survey was designed to be completed within 20-30 minutes. Written informed consent was obtained from all individual participants included in the study. Irrespective of survey completion, participants were eligible to enter a draw for a chance to win a $50 CAD Amazon gift card. The study was approved by The University of British Columbia (Canada) Behavioural Research Ethics Board (ID: H20-01855).

### Recruitment and Eligibility

#### Recruitment

Registered dietitians specializing in MBS throughout Canada were identified using snowball and convenience sampling. They were contacted and asked to disseminate the study information (such as recruitment posters) to their patients. Advertisements through Obesity Canada social media network were also used to recruit participants. The recruitment spanned from June 30, 2020 to April 30, 2021.

#### Eligibility

Eligible participants included 1) individuals who have undergone MBS, 2) caregivers of individuals who have undergone MBS or, 3) healthcare providers (such as registered dietitians) specializing in MBS. Participants were excluded if they did not meet the eligibility criteria or if they could not read in English or French and lacked access to a device with Internet connection.

### Development of the nutrition education tool: *Protein Cards*

The development of *Protein Cards* was inspired by the dietitians at the Centre of Excellence in Adolescence Severe Obesity (CEASO, Montreal Children’s Hospital, Montreal, QC) and co-author Cohen. The tool is available in both English and French and includes 40 easy-to-follow recipes and food items, which were color coded to help guide patients meet their protein requirements for the various post-MBS diet stages: fluid(blue), purée (green), soft (pink), and regular (yellow). This tool focuses on recipes that are protein-rich, with a novel method of quantifying protein. Protein content was denoted in 10 g increments adapted from the carbohydrate counting methodology [42] in the form of a fish icon, which aims to simplify monitoring protein intake without complex calculations. The cards (n= 84) are double-sided: the front displayed the color-coded diet stage, an image of the completed recipe/food item, and protein content per serving while the back provided the recipe or food item along with tips for food substitutions, flavor enhancements, preparation, information on where to source ingredients, and food storage (S1 Fig).

### Procedures

The self-administered survey was hosted through Qualtrics (Qualtrics International Inc., Seattle, Washington and Provo, Utah, United States), an online survey tool used for academic research at The University of British Columbia (Vancouver, British Columbia, Canada). The survey consisted of two parts: the first section included sociodemographic questions, and the second section utilized an adapted version of the Suitability Assessment of Materials (SAM) questionnaire to evaluate the content and suitability of chosen pages of the *Protein Cards*, which included the cover page, instruction page, and four representative recipes/ food items corresponding to each diet stage post-surgery (S1 Fig).

## Measures

### Suitability Assessment of Materials (SAM)

The suitability of *Protein Cards* was evaluated using SAM, a widely used and validated rating tool to assess the appropriateness of health information materials [43–46]. From the SAM questionnaire, a total of 21 questions were carefully selected and adapted to modify the wording to accommodate literacy level of all ages. These questions were classified into six categories: content, literacy demand, graphic illustrations, layout and typography, learning stimulation and motivation, and cultural appropriateness. An additional two questions were included to understand the usability of the cards [how likely would you consult the recipe book during the following periods: fluid, purée, soft and regular diet?] and to determine the preferred delivery format [how likely would you use the recipe book in the following formats: paper book, electronic book, mobile application, and webpage?].

Participants were asked to assess the *Protein Cards* using three possible ratings for each question: “superior”, “adequate”, or “not suitable”, which were assigned point scores of 2, 1, and 0, respectively. The total SAM score was calculated as the sum of earned points divided by the total maximum score of 42. The final SAM scores were reported as percentages, with 70-100% indicating superior material, 40-69% adequate material, and 0-39% not suitable material. Participants rated the usability of the cards during the various diet stages and the preferred delivery format on a scale of 0 to 100 (0 = not at all; 100 = highly).

### Data Analysis

Quantitative survey results were obtained in TSV format, and all analyses were done with R version 4.3.1 (The R Foundation for Statistical Computing, Vienna, Austria). Descriptive statistical analysis was conducted to report characteristics of the participants as total counts and percentages of the total. SAM assessment scores were also calculated as total counts and reported as percentages for the different SAM categories. The summary statistics of the likelihood of *Protein Cards* use across diet stages of MBS and its preferred method of delivery were reported as means and standard deviations, wherever appropriate. This analysis was repeated independently for patients, healthcare providers, and caregivers, to gain a better understanding of the participants’ preferences.

## Results

### Participant Characteristics

Of the 1,836 surveys received, 442 survey responses were included in the analysis. Participants were excluded if they did not meet inclusion criteria (n = 237), absence of consent (n = 1), missing/ incomplete surveys (n= 177) and data validity, including recurring email and IP addresses (n = 842), inappropriate email address (n = 1), duplicated written responses (n= 33), and completion time under five minutes (n = 103) (Fig 1). Among the 442 participants, 268 (60.63%) were individuals who have undergone MBS, 106 (23.98%) were healthcare providers specializing in MBS, and 68 (15.38%) were caregivers of patients who had MBS. Majority of participants were female (62.67%) aged between 19-39 (80.00%), self-identified as White (59.18%), completed college/ university educational level (31.30%) and reported an individual annual income of $45,000-$74,999 CAD (43.66%) (Table 1).

**Fig 1.**
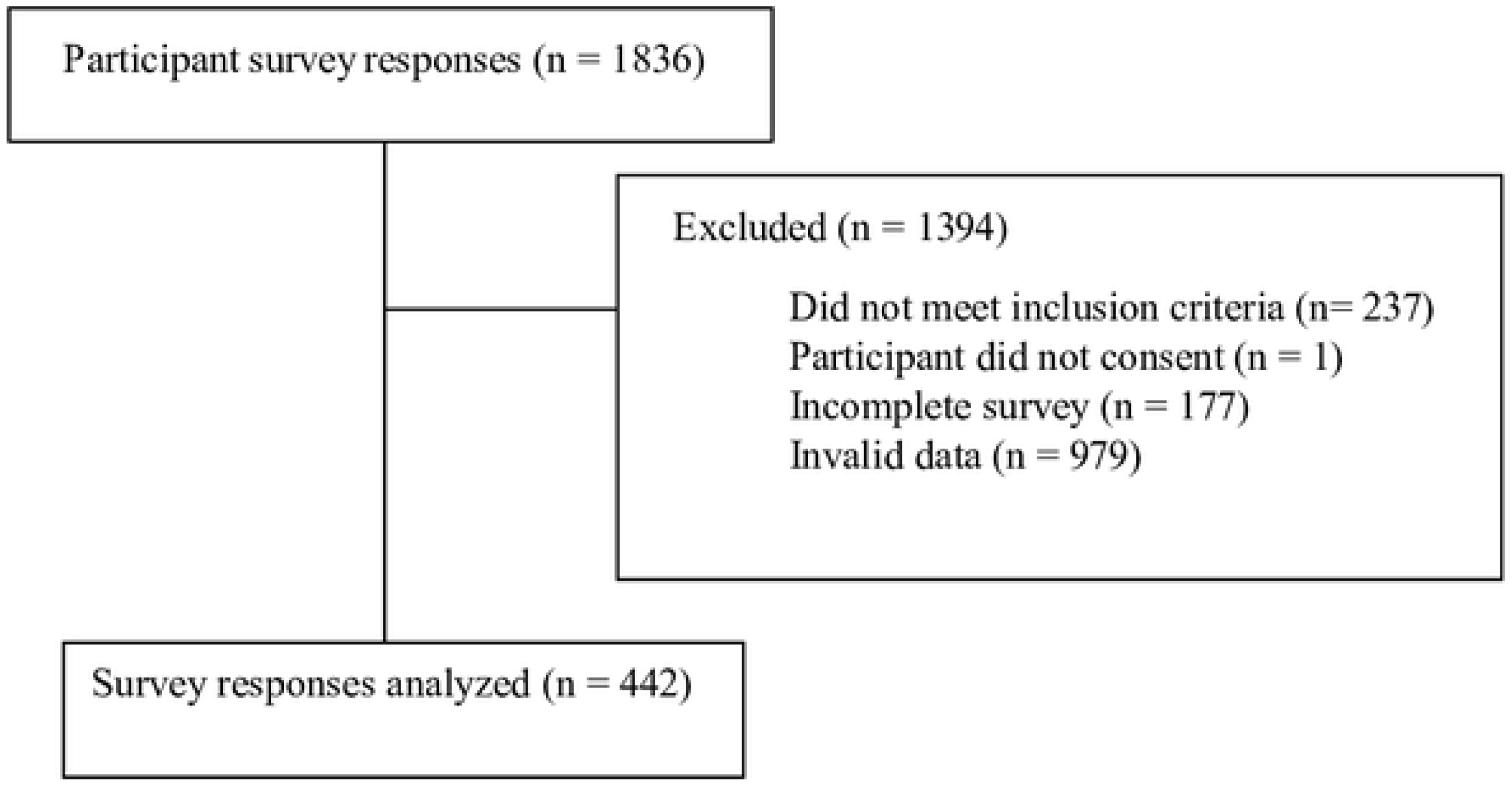
Adapted CONSORT Diagrant for Participant Flow.

**Table 1.** Characteristics of Survey Participants, 𝒏 = 𝟒𝟒𝟐.

| Characteristics | Count (%) |
| --- | --- |
| Age |  |
| 14-18 | 5 (1.14) |
| 19-29 | 154 (35.00) |
| 30-39 | 198 (45.00) |
| 40-49 | 57 (12.95) |
| 50-59 | 24 (5.45) |
| 60-69 | 2 (0.45) |
| Not specified | 2 (0.45) |
| Sex |  |
| Male | 165 (37.33) |
| Female | 277 (62.67) |
| Ethnicity |  |
| Aboriginal/ First Nations/ Métis | 65 (14.74) |
| Arab | 2 (0.46) |
| Black | 48 (10.89) |
| Asian |  |
| Chinese | 11 (2.50) |
| Filipino | 2 (0.45) |
| South Asian (East Indian, Pakistani, Sri Lankan, etc.) | 24 (5.44) |
| West Asian (Iranian, Afghan, etc.) | 1 (0.23) |
| Latin American | 9 (2.04) |
| White | 261 (59.18) |
| Mixed | 18 (4.07) |
| Not specified | 1 (---) |
| Occupation |  |
| Patients | 268 (60.63) |
| Dietitians | 78 (17.65) |
| Healthcare providers | 28 (6.33) |
| Caregivers | 68 (15.38) |
| Education level |  |
| Elementary school | 26 (5.90) |
| High school | 49 (11.11) |
| CEGEP | 209 (47.39) |
| Vocational school or apprenticeship training | 18 (4.08) |
| College | 75 (17.01) |
| University | 63 (14.29) |
| Prefer to not answer | 1 (0.23) |
| Not applicable | 1 (---) |
| Income |  |
| Less than \$15,000 | 29 (6.56) |
| \$15,000 - \$29,000 | 58 (13.12) |
| \$29,000 - \$44,999 | 74 (16.74) |
| \$45,000 - \$59,999 | 100 (22.62) |
| \$60,000 - \$74,999 | 93 (21.04) |
| \$75,000 or more | 82 (18.55) |
| Prefer to not answer | 6 (1.36) |
| Not specified | --- |

### SAM Questionnaire

The *Protein Cards* tool was scored as a “superior” postoperative nutrition education tool for MBS, with an overall rating of 73.16%. Each of the six SAM categories received superior mean score values: content (74.2%), literacy demand (72.1%), graphic illustrations (74.7%), layout and typography (71.9%), learning stimulation and motivation (73.6%), and cultural appropriateness (72.2%) (Table 2). When examining the data by participant groups, higher mean scores were rated for content by caregivers (76.47%), and cultural appropriateness by healthcare providers (71.32%) while patients rated the tool higher for literacy demand, graphic illustrations, learning stimulation and motivation (74.81%, 76.91%, 75.42%, respectively) (Table 3).

**Table 2.**
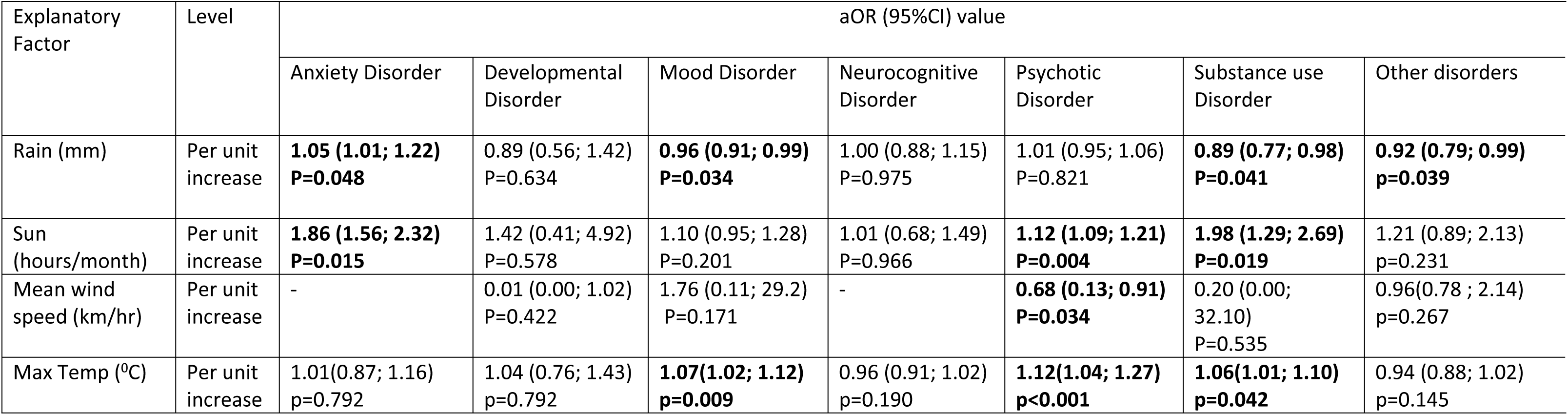
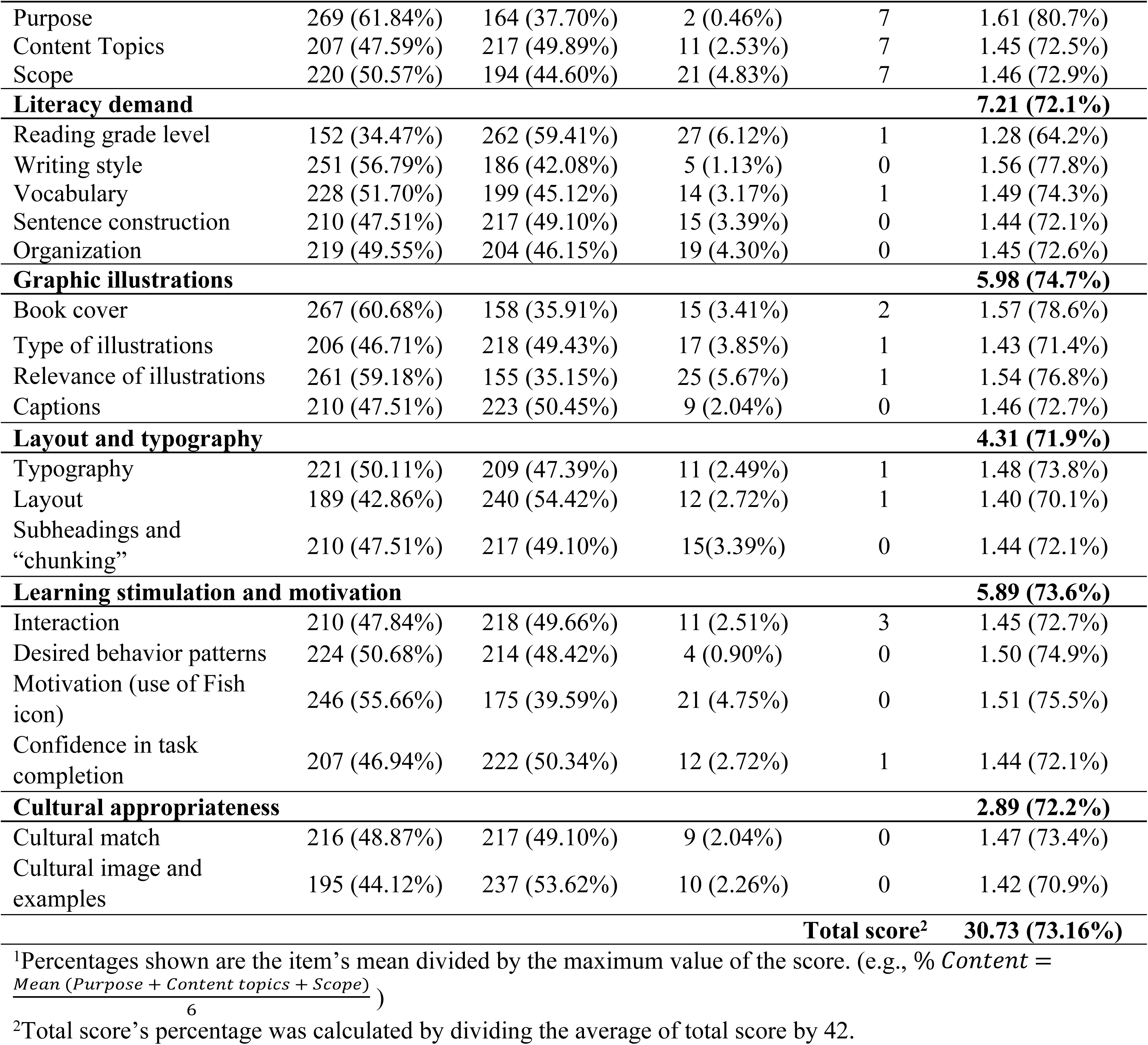
Suitability Assessment of Materials (SAM) Scores by Category.

**Table 3:** Overall SAM Scores among Participants.

| SAM scores by category (overall) | Count (%) of participants’ overall evaluation<sup>1</sup> |  |  | Mean score (%)<sup>2</sup> |
| --- | --- | --- | --- | --- |
|  | Not suitable | Adequate | Superior |  |
| Content |  |  |  |  |
| Patients | 9 (3.36) | 102 (38.06) | 157 (58.58) | 4.55 (75.81%) |
| Dietitians | 5 (6.41) | 42 (53.85) | 31 (19.74) | 4.08 (67.95%) |
| Healthcare providers | 1 (3.57) | 15 (53.57) | 12 (42.86) | 4.21 (70.24%) |
| Caregivers | 1 (1.47) | 29 (42.65) | 38 (55.88) | 4.59 (76.47%) |
| Literacy demand |  |  |  |  |
| Patients | 2 (0.75) | 70 (26.12) | 196 (73.13) | 7.48 (74.81%) |
| Dietitians | - | 36 (46.15) | 42 (53.85) | 6.68 (66.80%) |
| Healthcare providers | 1 (3.57) | 8 (28.57) | 19 (67.86) | 6.64 (66.43%) |
| Caregivers | - | 27 (39.71) | 41 (60.29) | 7.00 (70.00%) |
| Graphic and illustrations |  |  |  |  |
| Patients | 5 (1.87) | 64 (23.88) | 199 (74.25) | 6.15 (76.91%) |
| Dietitians | 7 (8.97) | 38 (35.90) | 43 (55.13) | 5.56 (69.55%) |
| Healthcare providers | 1 (3.57) | 14 (50.00) | 13 (46.43) | 5.54 (69.20%) |
| Caregivers | 3 (4.41) | 21 (30.88) | 44 (64.71) | 5.94 (74.27%) |
| Layout and typography |  |  |  |  |
| Patients | 3 (1.12) | 151 (56.34) | 114 (42.54) | 4.35 (73.51%) |
| Dietitians | 2 (2.56) | 51 (65.38) | 25 (32.05) | 4.08 (67.95%) |
| Healthcare providers | - | 20 (71.43) | 8 (28.57) | 3.96 (66.07%) |
| Caregivers | 3 (4.41) | 24 (35.29) | 41 (60.29) | 4.57 (76.23%) |
| Learning stimulation and motivation |  |  |  |  |
| Patients | 2 (0.75) | 80 (29.85) | 186 (69.40) | 6.03 (75.42%) |
| Dietitians | 4 (5.13) | 35 (44.87) | 39 (50.00) | 5.51 (68.91%) |
| Healthcare providers | 1 (3.57) | 10 (35.71) | 17 (60.71) | 5.64 (70.54%) |
| Caregivers | - | 22 (32.35) | 46 (67.65) | 5.85 (73.16%) |
| Cultural appropriateness |  |  |  |  |
| Patients | 4 (1.49) | 80 (29.85) | 184 (68.66) | 2.92 (72.95%) |
| Dietitians | 3 (3.85) | 24 (30.77) | 51 (65.38) | 2.80 (69.87%) |
| Healthcare providers | 1 (3.57) | 8 (28.57) | 19 (67.86) | 2.93 (73.21%) |
| Caregivers | 3 (4.41) | 20 (29.41) | 45 (66.18) | 2.85 (71.32%) |

<sup>1</sup>In the overall evaluation, 0–39% is considered ‘not suitable’, 40–69% is ‘adequate’ and 70% is ‘superior’. We showed the number of participants that falls under each category by dividing their overall score with the maximum score of the category.
<sup>2</sup>Mean score is the average of total score and the percentage was calculated by dividing the mean with the maximum score of the category.
Abbreviation used: SAM; Suitability Assessment of Materials

When participants were asked at which post-MBS stage would they likely consult *Protein Cards*, it was reported that ∼64% would use the cards for the soft diet stage (63.98 ± 20.71). This was followed by regular diet (63.68 ± 20.95), purée diet (58.58 ± 22.17), and fluid diet (57.54 ± 21.25) (Table 4). When considering the preferred delivery format of *Protein Cards*, mobile application (63.46 ± 20.44) was found to be the most favored option among caregivers (68.47 ± 19.51), healthcare providers (64.00 ± 23.93), patients (64.15 ± 19.25), and dietitians (56.44 ± 22.45). Other evaluated delivery formats included webpage (59.04 ± 22.77), paper book (59.42 ± 21.12), and electronic book (56.18 ± 20.94) (Table 5).

**Table 4.** Likelihood of *Protein Cards* Use Across Diet Progression Stages of Metabolic Bariatric Surgery among Patients, Healthcare Providers and Caregivers.

| **Diet Stage** | ***n*** | **Median** | **Mean ± SD** |
| --- | --- | --- | --- |
| **Full fluid diet** | 440 | 57.0 | 57.54 ± 21.25 |
| Patients | | 58.0 | 58.52 ± 21.64 |
| Healthcare providers |  |  |  |
| Dietitian | | 53.0 | 53.68 ± 20.76 |
| Others | | 58.0 | 58.57 ± 21.61 |
| Caregivers | | 59.5 | 57.66 ± 20.02 |
| **Purée diet** | 441 | 60.0 | 58.58 ± 22.17 |
| Patients | | 61.5 | 61.83 ± 20.70 |
| Healthcare providers |  |  |  |
| Dietitian | | 49.0 | 49.96 ± 22.68 |
| Others | | 60.5 | 59.11 ± 24.75 |
| Caregivers | | 55.0 | 55.32 ± 23.46 |
| **Soft diet** | 439 | 64.0 | 63.98 ± 20.71 |
| Patients | | 67.0 | 66.26 ± 19.36 |
| Healthcare providers |  |  |  |
| Dietitian | | 58.5 | 57.37 ± 21.32 |
| Others | | 61.0 | 61.64 ± 25.19 |
| Caregivers | | 63.0 | 63.41 ± 21.88 |
| **Regular diet** | 440 | 62.5 | 63.68 ± 20.95 |
| Patients | | 65.0 | 65.32 ± 19.10 |
| Healthcare providers |  |  |  |
| Dietitian | | 53.0 | 54.51 ± 23.95 |
| Others | | 64.5 | 64.04 ± 25.07 |
| Caregivers | | 70.0 | 67.50 ± 20.06 |

**Table 5.**
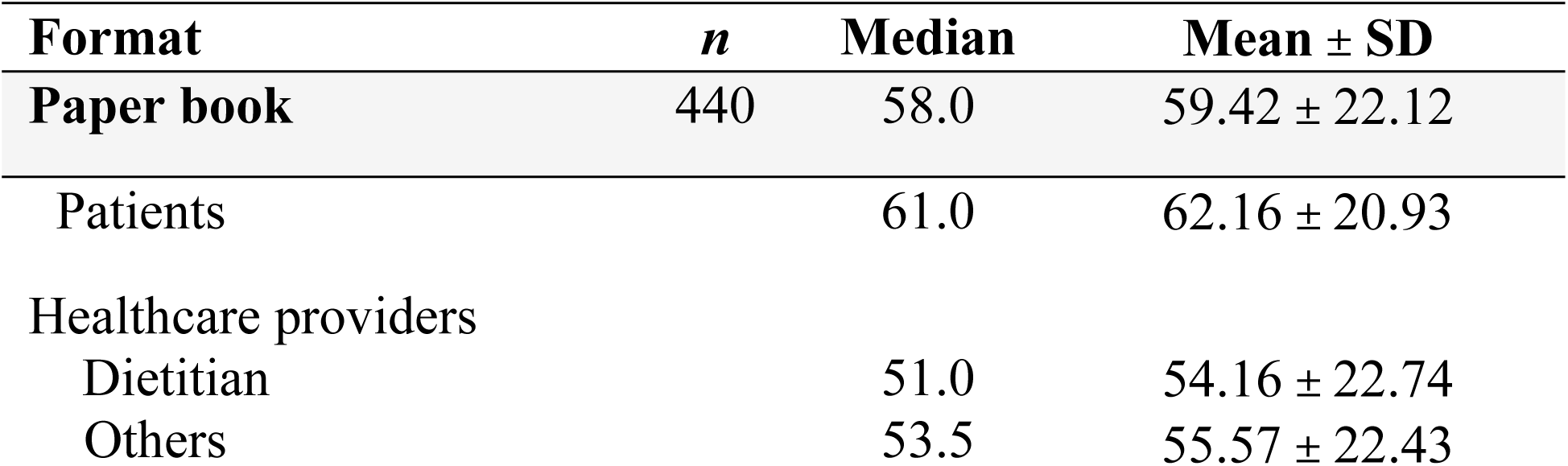
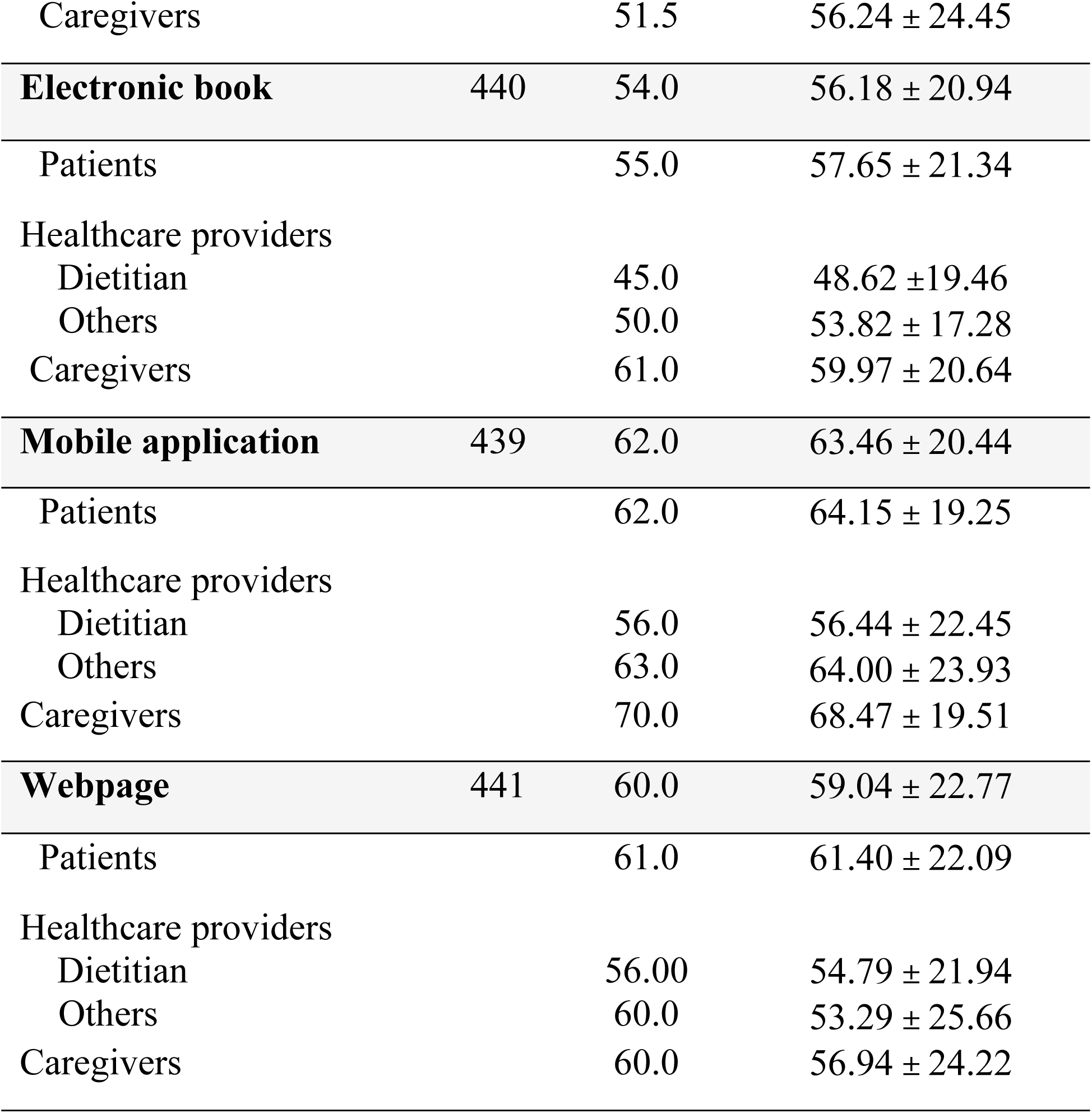
Preferred Delivery Format of *Protein Cards* among Patients, Healthcare Providers and Caregivers.

## Discussion

The current study assessed the suitability of *Protein Cards*, a postoperative patient nutrition education tool in a sample of individuals who have undergone MBS, their caregivers, and healthcare providers specializing in MBS. Research suggests that there is a lack of diversity of nutrition education materials tailored for the needs of individuals who undergo MBS [37,47] . The focus of *Protein Cards* is to provide easy-to-follow recipes that are protein-rich to facilitate healthy dietary habits and meet protein requirements during the post-MBS diet stages.

Creating suitable dietary education tools for the intended audience is critical to support the adoption of healthy dietary behavior change. Participants reported *Protein Cards* to be a “superior” tool with an overall rating of 73.16%, which aligns or exceeds the ratings of other studies that assessed health educational materials [44–46]. The *Protein Cards* received a superior rating for content-purpose (80.7%), indicating participants’ clear understanding of the tool’s objectives in addressing the dietary needs, particularly protein intake following MBS. Additionally, such a rating suggests the tool’s suitability in supporting optimal transition through the various diet stages and in facilitating the adoption of healthy dietary habits.

Different aspects of nutrition education resources need to be considered to ensure the tool’s suitability. One of those is the tool’s readability. In this study, participants rated the readability of *Protein Cards* to be between sixth to eighth grade reading level, which was likely due to the use of plain language. Other studies have assessed the readability of MBS patient materials and found them to be overall of poor quality; these materials [35,36,38,39] were above the recommended sixth grade reading level. In a specific examination of patient information publications from the American Society of Metabolic and Bariatric Surgery, Massie et al. (2024) utilized multiple validated assessment tools to evaluate the readability of the publications. Out of the n= 11 publications analyzed, none met the recommended sixth grade reading level, suggesting refinements are needed. Given these findings, there is significant opportunity to enhance the reading grade level of *Protein Cards*, ensuring the use of clear and plain language to better support patients in meeting their protein needs post-MBS.

Researchers who work in the area of developing nutrition education tools should consider the preferred delivery format to enhance accessibility and user satisfaction of these educational tools. The likelihood of participants utilizing the *Protein Cards* varied based on delivery format, including paper book, electronic book, mobile application, and webpage. Participants noted a high preference for a mobile application, which has been postulated to reach and gain more engagements among individuals. [48] This is consistent with findings of studies that explored adults’ perspectives on patient health education materials, which have shown a preference for varied types of electronic and paper resources[49,50]. This highlights that the *Protein Cards* might be more accessible for a diverse sample of patients if they are available in diverse delivery formats.

MBS is a complex process the extends beyond the surgery itself, involving not only the patients but also their healthcare providers and caregivers to ensure long-term success of dietary management. Our study addresses this by concurrently evaluating the suitability of a nutrition education tool among these groups, with the aim to refine the tool to make it more comprehensive, suitable, and practical for implementation in clinical practice. Research suggests that patients can meet nutritional needs with healthy food choices, [51] but experience challenges adhering to postoperative guidance. This suggests that lifelong support is needed, which may lead to improved self-efficacy, a hallmark for behavioral compliance. [52] Additionally, social support from family and friends has been shown to significantly influence dietary management, [52,53] physical and psychological well-being, [53,54] by facilitating effective coping strategies for stressors – a need that becomes more critical as patients adjust to their “new normal” post-MBS. [55] In a study that explored experiences of partners of individuals who have undergone MBS found that they faced various changes, including their level of engagement with the MBS process, such as efforts to access resources available to have a better understanding of the surgical process, and adoption to behavioral changes made by their spouses. [56] While another study highlighted that healthcare providers working in Canadian bariatric surgery program experienced challenges in making clinical decisions on how to best guide patients regarding nutrition pre- and post-surgery [57]. These demonstrated the ongoing need for bariatric resources that are user-friendly, easily accessible, and adaptable to different patient profiles. These findings convey the importance of a patient-centered approach that considers patients, their caregivers, and healthcare providers as integral stakeholders in MBS.

The strengths of this study include a large sample size, and the diverse engagement of stakeholders (patients, caregivers, and healthcare providers) provided a multifaceted evaluation of the *Protein Cards*. We also used SAM, an evaluation questionnaire that surveys on different aspects of patient education materials. However, this study has limitations, including the lack of data on the type of MBS, which could provide further insights into the tailored nutritional needs of patients.

In summary, there is a critical need for clear, accessible nutrition education tools tailored for MBS. Our findings demonstrate that the tool we have developed, *Protein Cards*, is usable and suggests potential benefits for meeting protein requirements across the various stages of post-MBS diet. Participants found the tool to be superior, with a high likelihood of use during these stages. Such tools are essential for supporting patients navigate this complex journey and are pivotal in advancing bariatric care. This can aid in facilitating a smoother transition to a broader range of foods and assist with the emotional and psychological challenges associated with dietary intake post-MBS. Future work will involve refining the tool and pilot testing the tool in clinical settings with individuals undergoing MBS, which will inform a future larger intervention trial aimed at evaluating the efficacy of *Protein Cards* in helping meet protein recommendations post-MBS.

## Data Availability

The data underlying the results presented in the study are available from the Nutrition and Eating Behaviour Lab website, https://nutritioneatingbehaviour.landfood.ubc.ca

## Acknowledgements

This study was supported by funding from Cohen’s University of British Columbia Start-Up Funds and Montreal Children’s Hospital Foundation. The authors wish to acknowledge all the individuals that contributed to this study: Yolanda Wang and Clare Douglas. Special thanks to Obesity Canada for their role in advertising the study.

## Supporting Information

**S1 Fig. Illustrative Pages of the Protein Cards.**

